# Clinical application of Complete Long Read genome sequencing identifies a 16kb intragenic duplication in EHMT1 in a patient with suspected Kleefstra syndrome

**DOI:** 10.1101/2024.03.28.24304304

**Authors:** John E. Gorzynski, Shruti Marwaha, Chloe Reuter, Tanner D. Jensen, Alexis Ferrasse, Archana Natarajan Raja, Liliana Fernandez, Elijah Kravets, Jennefer Carter, Devon Bonner, Shirley Sutton, Undiagnosed Diseases Network, Maura Ruzhnikov, Louanne Hudgins, Paul G Fisher, Jonathan A. Bernstein, Matthew T. Wheeler, Euan A. Ashley

## Abstract

Long read sequencing offers benefits for the detection of structural variation in Mendelian disease. Here, we applied a new technology that generates contiguous long reads via tagmentation and sequencing by synthesis to a small cohort of patients with undiagnosed disease from the Undiagnosed Diseases Network. We first compare sequencing from the HG002 benchmark sample from Genome In A Bottle using nanopore sequencing (R10.4.1, duplex reads, Oxford Nanopore), single molecule real time sequencing (Revio SMRT cell, Pacific Biosciences) and complete long read sequencing (S4 flowcell, Novaseq, Illumina). Coverage was 33-35x across platforms. Read length N50 was 6.5kb (ICLR), 16.9kb (SMRT), and 33.8kb (ONT). We noted small differences in single nucleotide variant F1 scores across long read technologies with single nucleotide variant F1 scores (0.985-0.999) exceeding indel scores (0.78-0.99) and structural variant scores (0.74-0.96). We applied CLR sequencing to seven undiagnosed patients. In one patient, we detected and prioritized a novel 16kb intragenic duplication encompassing exons 5 and 6 in *EHMT1*. Resolution of the breakpoints and examination of flanking sequences revealed that the duplication was present in tandem and was predicted to result in a frameshift of the amino acid sequence and an early termination codon. It resulted in a diagnosis of Kleefstra syndrome. The variant was confirmed with targeted *EHMT1* clinical testing and detected via nanopore and SMRT sequencing. In summary, we report the early clinical application of complete long read sequencing to a small cohort of undiagnosed patients.

## INTRODUCTION

Approaches to the diagnosis of rare Mendelian disease have advanced rapidly in recent years, yet are still largely reliant on exome and genome sequencing (Wojcik et al. 2023). These short read-based sequencing technologies have been adopted as gold standard due to their high sequencing quality, and have a diagnostic rate ranging from 20-50% (Yang et al. 2014; Splinter et al. 2018; Cohen et al. 2022; Lionel et al. 2018). However, the advent of long read sequencing technologies has challenged the standard of short read sequencing based genomic testing for rare disease. Whereas short read sequencing produces reads ranging in size of 150-300 nucleotides, long read sequencing produces kilobase to megabase reads (Logsdon et al. 2020). In turn, long read sequencing offers improved mapping to highly homologous, repetitive or GC rich regions of the genome (van Dijk et al. 2018; Chintalaphani et al. 2021). Hard to characterize variants, such as large insertions, deletions, translocations, and tandem repeats, are more readily detected with long read sequencing (Ebert et al. 2021; Mastrorosa et al. 2023). As such, patients with Mendelian diseases may benefit from more comprehensive genomic analyses harnessing long read sequencing (Chintalaphani et al. 2021; van Dijk et al. 2018).

Three long read sequencing technologies include: nanopore sequencing (“ONT”, Oxford Nanopore Technologies, Oxford UK), Single-Molecule, Real-Time (SMRT) sequencing (“PacBio” Pacific BioSciences, Menlo Park CA), and Illumina Complete Long Read sequencing (“ICLR” Illumina inc. San Diego CA). The newest ICLR sequencing technology developed by Illumina can produce long read data with read length N50’s ranging from 5-7 kilobases (kb). A 1-2 day library preparation protocol requires as little as 10 nanograms of input genomic DNA and begins with an initial tagmentation step which fragments the DNA to approximately 10kb. Each fragment is then ‘land-marked’ i.e., enzymatically marked with unique patterns of single base pair changes at a frequency of 5-7%. The DNA molecules, now containing a unique land-mark profile, are amplified and exposed to another round of tagmentation that facilitates sequencing on a standard illumina flow cell in the Novaseq system. In addition to the ICLR library, a standard unmarked library must be prepared for 30X short read sequencing by synthesis for the computational pipeline as seen below. Bioinformatics analysis pipeline involves generation of complete long reads and their integration with short read, unmarked WGS library with 30x coverage from the same sample to produce high-quality variant calls. The first step in this workflow is identification of reads with shared land-marks, which involves construction of a network of shared marks. The strength of the connection between reads depends on the number of shared marks and the number of conflicting marks. The network of connected reads is decomposed into clusters of strongly-connected reads. Each cluster corresponds to a set of reads putatively coming from a single template molecule, assembled into a land-marked long read. After the long reads are generated, the marks are removed. Each assembled land-marked long read is compared to unmarked reads to differentiate land-marks from true variants. Land-marked bases that conflict with unmarked reads are updated to match the unmarked reads. This is followed by secondary analysis where long and short reads are aligned and called separately. The variants from both short and long read are later merged and processed to generate a single variant file, one for SNVs/Indels and one for structural variants(Roessler).

In this work, we describe a comparative analysis of these three long read sequencing technologies. We piloted the application of ICLR sequencing on a cohort of seven suspected Mendelian disease patients. We show that ICLR sequencing detects a large, diagnostic structural variant in a patient with *EHMT1* Kleefstra syndrome. This is to our knowledge the first report of the application of ICLR sequencing in human rare disease diagnostics.

## RESULTS

### Comparison of long read technologies using HG002

We acquired publicly available data for Genome In A Bottle sample, HG002 (Zook et al. 2016), generated by all of the commercially available long read sequencing technologies. Each data set was generated using the latest technology from each sequencing company. ICLR was prepared with the standard Illumina complete long read kit and sequenced on an S4 flowcell, and was coupled with short read (Sequencing by synthesis (SBS)) data generated with an Illumina PCR free prep sequenced on an S4 flowcell. Nanopore sequencing libraries were prepared with a ligation based library prep and sequenced on a R10.4.1 with duplex reads. SMRT sequencing libraries were prepared with a SMRTbell prep kit 3.0, and sequenced on a Revio SMRT Cell. Each sequencing data set was aligned to both GRCh37 and GRCh38, and variants were called (see data processing section in methods for variant calling details).

HG002 was sequenced at comparable 33-35x coverage across all three technologies, and we calculated a number of standard sequencing metrics including read length, and base calling quality scores (Table 1). The largest median read length (33.8kb) and read length N50 (22.9kb) was observed in the nanopore sequencing data. These longer reads allowed phasing of more variants and achieved maximum phase blocks exceeding 12mb when compared to the other technologies (Supplemental Figure 1). The highest median quality score (30), and percent identity (99.9%) was observed in the ICLR data.

**Table 1.**
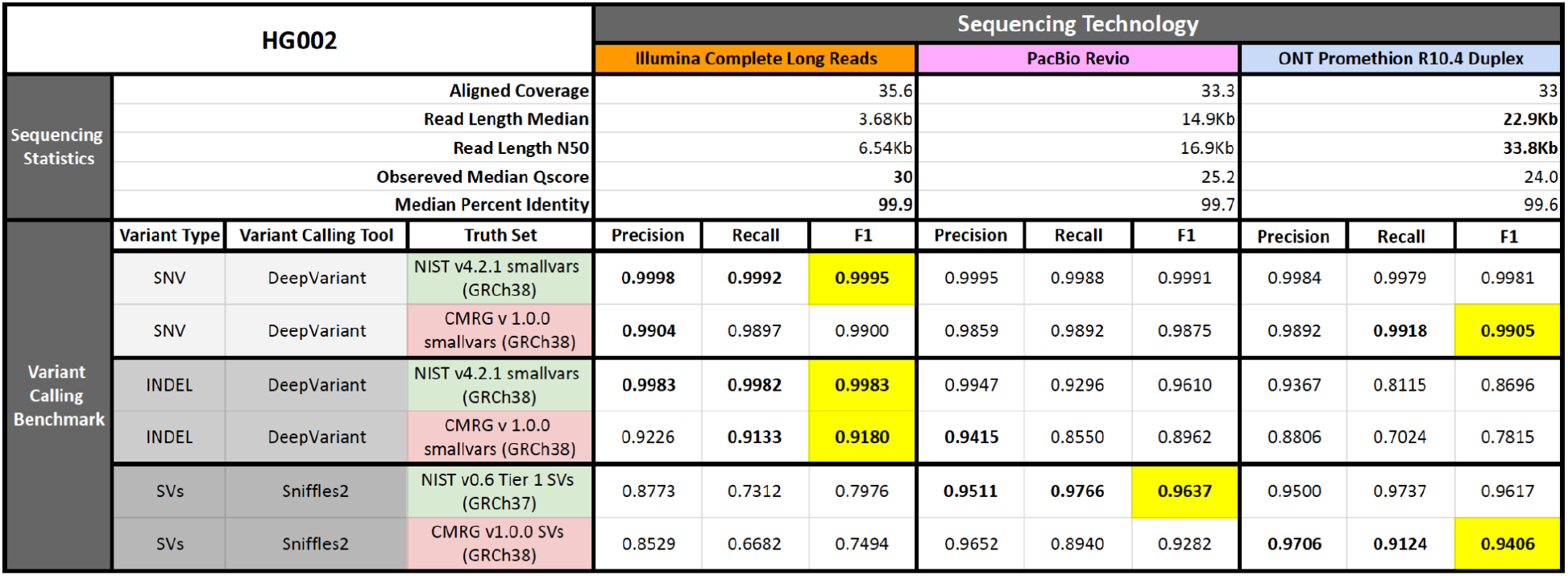
Sequencing a standard sample using three long read technologies: Illumina’s Complete Long Read (ICLR),nanopore sequencing: Oxford Nanopore Technologies (ONT) and Single Molecule Real-Time sequencing (SMRT): Pacific Biosciences (PacBio).

We compared variant call files (VCFs) from the three technologies against known SNV, indel, and SVs using the following truth sets: NIST v4.2.1 smallvars, CMRG v 1.0.0 smallvars, NIST v0.6 Tier 1 SVs, CMRG v1.0.0 SVs (Table 1, Supplemental Figure 1). The NIST data sets encompass genome wide tier 1 regions, whereas the CMRG data sets include medically relevant genes in regions of the genome known to be difficult to sequence. Marginal differences were observed in single nucleotide variants (SNV) F1 scores (range 0.9875-0.9995), however ICLR and nanopore performed best in the NIST smallvars and CMRG smallvars datasets, respectively. ICLR and SMRT achieved the highest indel F1 scores in the NIST and CMRG datasets respectively. SMRT and nanopore sequencing had the highest precision and recall in the NIST SV and CMRG SV datasets respectively.

Next we compared the coverage of approximately 4600 challenging medically relevant genes (CMRG’s) by the three long read technologies (Figure 1). At least 10x coverage over exonic bases was observed in approximately 4500 CMRG’s using the ICLR and SMRT, and over 4580 using nanopore sequencing. Nanopore performs the best due to its long N50. Although the coverage for ICLR and SMRT look comparable in this region, it should be noted that ICLR median coverage across these CMRG exons is 39x, whereas it is 32x for PacBio.

**Figure 1.**
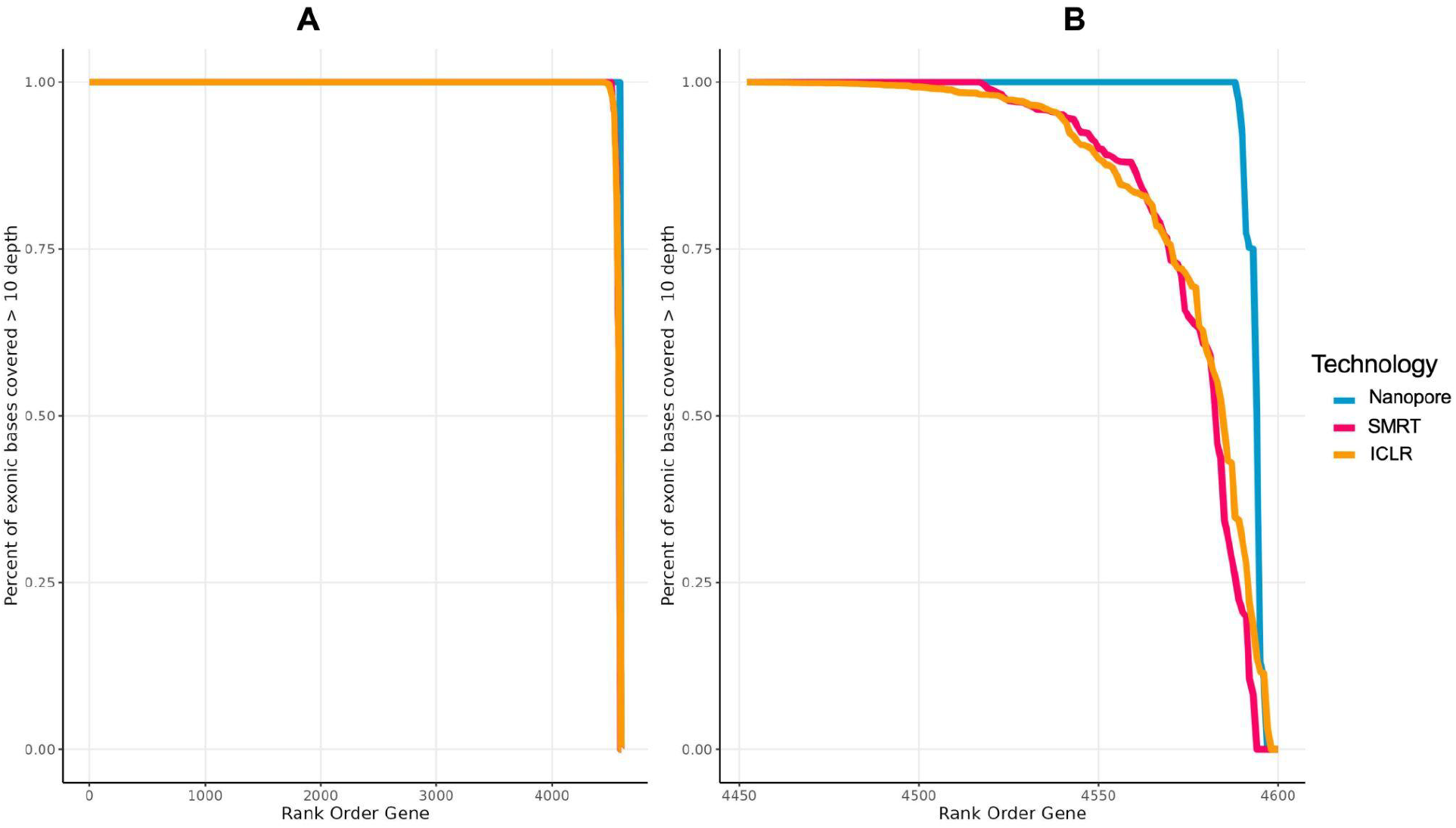
(A) Coverage of about 4600 challenging medically relevant genes (exons) by the three long read technologies. (B) is a zoomed view on the tail end genes where there is less than 100% exonic coverage in any of the technologies.

### ICLR sequencing of a rare disease cohort

To explore the possibility that ICLR sequencing could detect a causal variant not identified with shortread genome sequencing, we next performed ICLR sequencing on a cohort of seven participants from the Stanford Center for Undiagnosed Diseases with suspected Mendelian disease in addition to HG002 as an internal control (Table 2). We generated an average of 108 gigabases of ICLR data that mapped to GRCh38, which resulted in a mean coverage of 34x (Supplemental Figure 2). When combined with short read sequencing by synthesis (SBS) data the mean coverage was 80.77. The median N50 value for ICLR bams across seven participants was 6.625 kb (Figure 2). We observed more SVs in the ICLR data when compared to the SBS alone.

**Figure 2.**
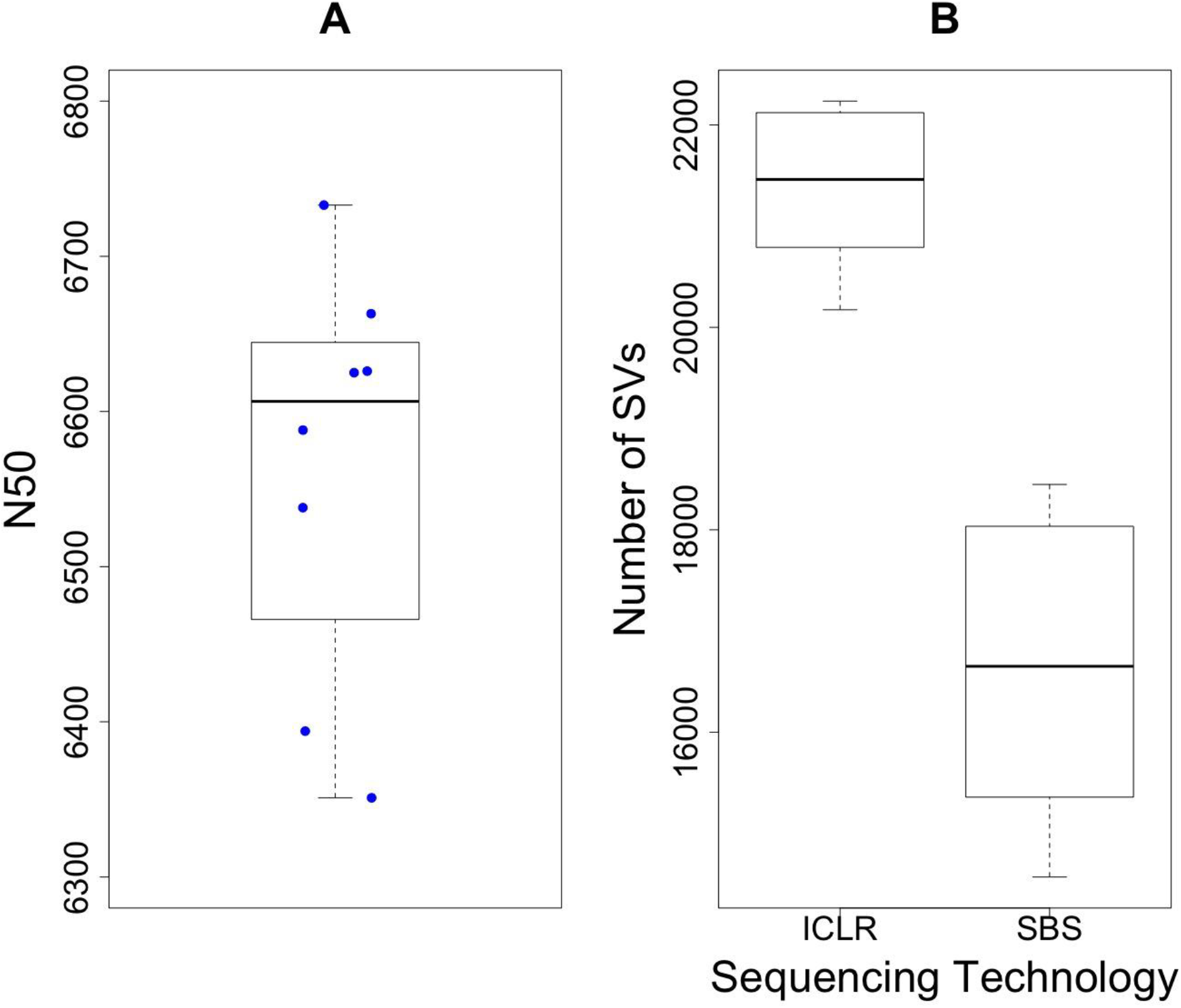
(A) Distribution of N50 from ICLR sequencing on seven patients with suspected rare diseases and HG002 sample. Each dot represents an individual sample. (B) Number of structural variants (SVs) called on the ICLR and SBS technologies.

**Table 2.** Clinical characteristics of rare disease participants that had ICLR sequencing

| Participant ID | Sex | Age (range, years) | Human Phenotype Ontology (HPO) terms |
| --- | --- | --- | --- |
| P1 | F | 51-55 | Progressive spastic paraparesis, Spasticity, Gastroesophageal reflux, Urinary urgency, Diplopia |
| P2 | M | 51-55 | Distal muscle weakness, Tongue fasciculations, Distal sensory impairment, Elevated circulating creatine kinase concentration, Muscle spasm, Lower limb dysmetria, Fatigue, Cough |
| P3 | F | 6-10 | Delayed speech and language development, Dystonia, Torticollis, Lower limb pain, Branchial cyst, Headache, Abdominal pain |
| P4 | F | 16-20 | Mild Intellectual disability, Global developmental delay, Delayed speech and language development, Developmental regression, Generalized hypotonia, Horseshoe kidney, Recurrent infections, Elevated circulating creatine kinase concentration, Paresthesia, Lower limb pain, Tip-toe gait, Otitis externa, Premature birth, Strabismus, Anxiety, Attention deficit hyperactivity disorder, Constipation, Migraine, Fine hair |
| P5 | F | 11-15 | Microcephaly, Mild conductive hearing impairment, Generalized limb muscle atrophy, Short stature, Bicuspid aortic valve, Neonatal respiratory distress, Micrognathia, Facial asymmetry, Low-set ears, Webbed neck, Down slanted palpebral fissures, Ptosis, Nasal speech, Talipes equinovarus, Achilles tendon contracture, Kyphoscoliosis, Knee flexion contracture, Camptodactyly, High palate, Keratosis pilaris, Fetal cystic hygroma, |
| P6 | F | 11-15 | Global developmental delay, Macrocephaly, Diffuse hypotonia, Autism spectrum disorder, Cataracts |
| P7 | M | 26-30 | Global developmental delay, Mild Intellectual disability, Infantile muscular hypotonia, Inguinal hernia, Hearing impairment, Overfolded helix, Hypermetropia, Synophrys, Delayed eruption of permanent teeth, Enuresis, Cutis marmorata, Clinodactyly, 2-3 toe syndactyly, Migraine, Highly arched eyebrow, Supernumerary nipple, Thoracic kyphosis, Pain insensitivity, Varicocele, Atrial fibrillation |
| HG002 |  |  | Genome in a Bottle Sample |

### Structural variant prioritization and interpretation

To assess for potentially pathogenic structural variants (SV) in each of the seven UDN participants, we employed our in-house SV prioritization pipeline that implements AnnotSV(Geoffroy et al. 2018) and StrVCTRE(Sharo et al. 2022) to generate a ranked list of possibly causal structural variants (Supplemental Methods). Review of the prioritized SVs for Participant 7 showed a duplication in the EHMT1 gene as the 2nd ranked candidate variant with an AnnotSV score of 1.1 and a StrVCTVRE score of 0.87. The prioritized EHMT1 SV was a 16kb intragenic duplication that encompasses exons 5 and 6 in the EHMT1 gene (NM_024757.4). Resolution of the breakpoints and examination of flanking sequences with ICLR sequencing visualization revealed that the duplication was present in tandem (Figure 3). It is predicted to result in a frameshift of the amino acid sequence and an early termination codon. The predicted impact of the intragenic duplication is supportive of a loss of function mechanism via haploinsufficiency of EHMT1. No equivalent duplications are present in the DECIPHER or gnomAD SV database. Comparing the clinical presentation of our patient to those described in typical Kleefstra syndrome patients we determined that this variant was diagnostic for Participant 7 (Supplemental Table 3). We reviewed other available long read sequencing data available on Participant 7 and found that the 16kb duplication within EHMT1 could also be appreciated with nanopore and SMRT sequencing. The duplication was clinically confirmed at a commercial laboratory. No diagnoses were made with ICLR sequencing in the remaining UDN participants.

**Figure 3.**
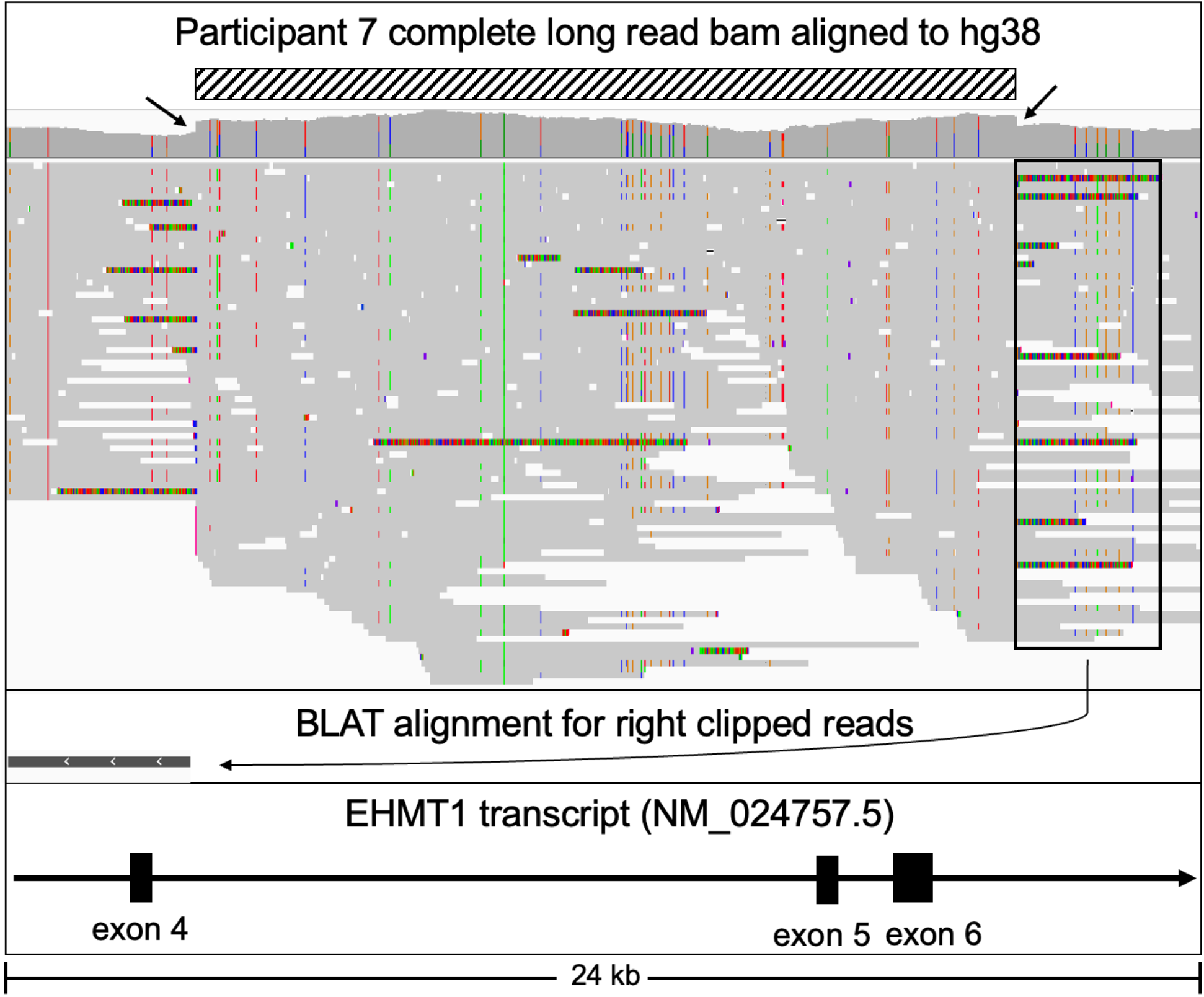
IGV visualization of ICLR sequencing encompassing a causal 16kb duplication event in the *EHMT1* gene. Striped bar indicates the region of the 16 kb duplication. Arrows in coverage histogram designate increase in height of coverage indicating duplication breakpoints. Top track = Participant 7 bam. Middle track = right clipped BLAT results for a read within the black rectangle supporting the tandem nature of duplication. Bottom track = Refseq transcript for EHMT1.

## DISCUSSION

We compare sequencing from two established long read sequencing technologies (nanopore and SMRT) and one recently released long read sequencing technology, Illumina Complete Long Read (ICLR). We further describe the clinical application of ICLR on patients with suspected Mendelian conditions. We successfully detected a diagnostic structural variant with these three long read technologies in a patient with Kleefstra syndrome.

The results of our comparative analysis of three long read sequencing technologies highlighted strengths and limitations of each. All three long read sequencing technologies identified SNVs with high precision and recall and there was marginal difference in their F1 scores. Early drawbacks of long read technologies included base calling errors (Ip et al. 2015; Laehnemann et al. 2016). However, long read technologies have continued to make improvements to their chemistry and tooling such that base calling is now on par with short read sequencing (Wang et al. 2021; Gorzynski et al. 2022; Wenger et al. 2019; Shafin et al. 2021; Wang et al. 2023). The ICLR sequencing technology was able to identify indels with the highest confidence which may be due to its reliance on complementary short read sequencing data in the data processing step. Throughout its historical development, nanopore sequencing has encountered challenges associated with small insertions and deletions (indels) (Goodwin et al. 2016), and our data suggests this remains a limitation most notably in homopolymer regions. We observed nanopore and SMRT sequencing data to have the highest performance in identification of structural variants, which is due to the larger read length N50s achieved by these platforms. Longer reads enhance mappability to the reference genome especially in hard to sequence regions of the genome, and therefore increases their ability to call structural variants. This is highlighted by nanopore read length N50 which is 2x larger than SMRT data and out performs all the technologies in the Challenging Medically Relevant Genes structural variant calls. Our analysis suggests that each long read sequencing technology has its strengths and limitations. These should be considered when selecting a technology to underlie a research or clinical investigation of the genome using long read sequencing.

As genomic technology advances, its clinical interpretation is key. We used a combination of annotation and prioritization tools to review variants called with the ICLR sequencing pipeline, which resulted in the curation of a likely pathogenic structural variant in a rare disease participant. The identification of a novel likely pathogenic *EHMT1* duplication expands our understanding of the disease-causing variant landscape of Kleefstra syndrome. Loss of function variants are well known to cause Kleefstra syndrome (Kleefstra et al. 2006, 2009). This variant is one of the smallest intragenic duplications reported to date across the literature and ClinVar (Schwaibold et al. 2014; Bonati et al. 2019; Yatsenko et al. 2012; Landrum et al. 2018). Representing a class of variant that has been historically difficult to detect, it is too small for microarray-based testing, even as short-read sequencing technology advances in copy number detection (Gross et al. 2019; Ho et al. 2020). This case highlights the value of long read sequencing, since the variant was not reported on clinical short read genome sequencing.

Thoughtful utilization of diverse genomic technologies is needed to maximize molecular diagnoses. There is not a universally acceptable approach to identifying genetic diagnoses and deciding on a combination of tests can be confusing for researchers and clinicians (Wojcik et al. 2023). The ICLR sequencing technology shortens the diagnostic gap left after short read genome sequencing technologies without the need to access additional hardware. Well known challenges of long read sequencing technologies in clinical settings are consistency of quality control, lack of universal implementation protocols, and immature bioinformatic tooling, and cost(Oehler et al. 2023). Compared to nanopore and SMRT sequencing technologies, ICLR sequencing technology does not require an additional sequencing machine beyond what is needed for short read sequencing. However ICLR sequencing does require short read sequencing in addition to the long read sequencing, increasing the time and cost of sequencing significantly.

We acknowledge two limitations of this study. The first limitation is the small sample size of Mendelian disease patients in our pilot study. Due to this, caution should be used when extrapolating the results to broader populations. The second limitation is that the ICLR data processing and analysis requires short read sequencing files to generate variant calls for a given sample, compared to ONT and PacBio which do not require any short read sequencing data for analysis. While ultimately, this results in the ICLR technology to more confidently identify SNVs and indels, it may be viewed as an unfair comparison to the other long-read technologies due to this requirement.

In summary, we describe the first human application of the new complete long read technology on patients with suspected Mendelian conditions. We successfully detected a diagnostic structural variant with all three on market long read sequencing technologies in a patient with Kleefstra syndrome.

## METHODS

### Ethics statement

Participants sequenced in this study were enrolled in the Undiagnosed Diseases Network (UDN) at Stanford Medicine and provided informed consent. The study is approved by the central IRB at the National Institutes of Health. Benchmarking data was generated from HG002 Genome in a Bottle sample that is publicly available from multiple sources (see Data Acquisition section).

### Rare disease cohort participant selection

We selected individuals enrolled in the Undiagnosed Diseases Network (CITE) at the Stanford Medicine clinical site to undergo ICLR sequencing. UDN participants have objective clinical findings in the absence of a unifying molecular diagnosis. Participants were prioritized for ICLR sequencing if we suspected they had a structural variant, nondiagnostic clinical short read genome sequencing, and available biological samples.

### HG002 Comparison Data Acquisition, Processing, and Variant Calling

HG002 for each of the long read sequencing technologies was acquired from:

<u>HG002 data sets:</u>

#### ICLR

HG002 Illumina complete long read data with incorporated sequencing by synthesis data was obtained from the illumina Basespace data repository (https://basespace.illumina.com/datacentral).

#### PacBio

HG002 data sequenced using the Pacific Biosciences Revio system was obtained from the PacBio public database. (https://downloads.pacbcloud.com/public/revio/2022Q4/HG002-rep1/)

#### Oxford Nanopore Technologies

Oxford Nanopore Duplexed reads from Genome in a Bottle sample HG002 was obtained from the Human Pangenome Project database. (https://human-pangenomics.s3.amazonaws.com/index.html?prefix=submissions/0CB931D5-AE0C-4187-8BD8-B3A9C9BFDADE--UCSC_HG002_R1041_Duplex_Dorado/Dorado_v0.1.1/stereo_duplex/)

SMRT and nanopore data were aligned to both GRCh37 and GRCh38 using Minimap2 (Li 2018, 2021). For the identification of small variants, including Single Nucleotide Variants (SNVs) and small insertions and deletions (INDELs), DeepVariant (Version 1.6.0) was employed(Poplin et al. 2018). To identify the best method for structural variant calling we benchmarked Sniffles2 (Smolka et al. 2024)against three other commonly used SV callers; cuteSV (2.1.0)(Jiang et al. 2020), SVIM (2.0.0)(Heller and Vingron 2019), and pbsv (2.9.0), as well as a consensus svVCF where independent calls from Sniffles2, cuteSV, and SVIM were merged using JasmineSV (1.1.5), insertions further refined with IrisSV(1.0.4), and variants detected by two or more SV calling tools were incorporated into the consensus VCF. Ultimately, Sniffles2 performed the best, and served as the basis for subsequent SV analysis.

ICLR data was processed, aligned to GRCh37 and GRCh38, and variant called using the DRAGEN ICLR WGS pipeline app, version 2.0.6 on Illumina basespace.

To benchmark variant calls detected by the three technologies, we downloaded relevant benchmarks from the National Institutes of Standards and Technologies for the Genome in a bottle HG002 truth set. These benchmarks covered SNVs, small indels, as well as deletion and insertion structural variants. For small variants, we use NIST v4.2 (GRCh38) truth set covering confident regions of the genome downloaded from (https://ftp-trace.ncbi.nlm.nih.gov/ReferenceSamples/giab/release/AshkenazimTrio/HG002_NA24385_son/NISTv4.2.1/GRCh38/). For structural variants we use NIST v0.6 Tier 1 (GRCh37) as a truth set downloaded from (https://ftp-trace.ncbi.nlm.nih.gov/ReferenceSamples/giab/release/AshkenazimTrio/HG002_NA24385_son/NIST_SV_v0.6/. Chromosome 20 was used for benchmarking of small variant calls using the tool hap.py (v0.3.15) for each of the three callers to calculate precision, recall, and F1. For SVs, technology SVs were compared against the NIST v0.6 Tier 1 truthset using Truvari v0.4.1.9 in order to calculate precision, recall, and F1. We also sought to benchmark variant calls in difficult to sequence regions of the genome, so we used the challenging medical region genes v1.0.0 from NIST to benchmark small variants and indels, downloaded from (https://ftp-trace.ncbi.nlm.nih.gov/ReferenceSamples/giab/release/AshkenazimTrio/HG002_NA24385_son/CMRG_v1.00/GRCh38/). We provide the snakemake workflow to call small variants and SVs from the publically available HG002 data and to run the benchmarking commands at this link: (https://github.com/tjense25/HG002_SeqTech_Comparison).

### ICLR Sequencing, and Data Processing

#### DNA Extraction

From whole blood or cultured fibroblasts, genomic DNA was extracted using Qiagen DNeasy Blood and Tissue Kit, or PureGene Kit (Qiagen, Hilden Germany) according to the manufacturer’s protocol. DNA was quantified using Qubit High Sensitivity DNA (Thermo Fisher Scientific,Waltham, MA; Ref: Q32853)

#### ICLR Sequencing

Illumina libraries were prepared according to the Illumina Complete Long Read Prep protocol (Illumina, San Diego, CA, USA; Ref: 20089108) and the documentation associated (Ref: 200015544 v00 April 2023) starting with 10 ng to 300 ng of DNA per sample as input. The TapeStation Systems 4150 with High Sensitivity D5000 Kit (Agilent, Santa Clara, CA, USA), and Qubit High Sensitivity (Thermo Fisher Scientific,Waltham, MA; Ref: Q32853) were used for final quality and quantity control for each prepared library. The molarity was adjusted to 0.875 nM (NovaSeq 6000) according to the manufacturer recommendations. Sequencing was accomplished on a NovaSeq 6000 with a single sample per lane on an S4 flowcell using the NovaSeq Xp workflow (151 cycles per read (2 x 151)).

#### Short Read Sequencing by Synthesis

Clinical short read genome sequencing was performed in accordance with CLIA practices at Hudson Alpha or Baylor Genetics. Sequencing was performed on either Illumina HiSeq X or Illumina NovaSeq (Supplemental Table 3).

#### ICLR Data Processing

The process of landmarking DNA to achieve long-reads results in non-native nucleotide incorporation into genomic DNA fragments. To align and further analyze this data, long reads must be assembled and the (nucleotide) landmarks must be removed. This process is enabled by Illumina DRAGEN which first identifies landmarked sites by one of two processes. In regions of the genome where sequencing reads map with confidence, landmarks are identified through alignment to the reference genome followed by detection of nucleotide variation. In genomic regions where reads do not map with confidence, a k-mer based approach is taken to identify the landmarks. Once all landmarks have been identified they are built into a weighted network to identify groups of reads from the same template. Once landmark reads are grouped, using a k-mer-based, de Bruijn graph-like method to assemble and generate long reads. Removal of the landmarks relies on comparison of the newly generated long reads to its associated short read sequencing data. Variants that are identified in both long and short reads are considered true variants, and those identified in long read alone are considered landmarks and are then removed from the sequence.

### Quality metrics of ICLR rare disease cohort

We used N50, a read or contig-length distribution metric to assess the read-length quality. N50 is the length of the shortest read or contig length obtained when the cumulative length of the longest read or contig length equals 50% of the total read or assembly length(Kim and Kim 2022; [CSL STYLE ERROR: reference with no printed form.]). First, samtools stats command (samtools version 1.16.1+htslib-1.16)(Danecek et al. 2021) was run on each ICLR bam file to calculate the read length and its frequency. N50 values were then calculated by an inhouse R script (R version 4.2.0 (2022-04-22))(Team 2017) using the above distribution of read lengths. Samtools depth and samtools stats commands were used to calculate mean coverage and bases mapped respectively.

### Variant annotation and prioritization of SVs for ICLR rare disease cohort

Variant calling done by Illumina using their new pipeline for ICLR data as described above. Structural variant prioritization was performed using an internal pipeline that integrates AnnotSV (version 3.0.5)(Geoffroy et al. 2018) and StrVCTVRE(Sharo et al. 2022). Input to the pipeline included SV vcf and HPO terms. Prioritized SNV/INDELs (from Exomiser version v12.1.0(Sharo et al. 2022; Smedley et al. 2015)) were also provided as an input for the structural variant pipeline to identify compound heterozygous variants. Prioritized outputs were reviewed by a trained genomic curator (CR). The top 10 structural variants were reviewed for phenotypic overlap, predicted impact of the SV on gene function, zygosity fitting the expected inheritance pattern, and rarity in population databases (gnomAD SV 3.0 and DECIPHER).

### Clinical orthogonal validation of candidate variants

Clinical genetic testing to orthogonally confirm candidate variants identified was performed via next generation sequencing panel at a commercial laboratory (Invitae).

## Data Availability

The data on HG002 Genome in a Bottle sample is available publicly and referenced in the
Methods section. The Ilumina complete long read sequencing data will be available in dbGaP in
accordance with Undiagnosed Diseases Network data sharing policies. As data is deposited
from the Undiagnosed Disease Network Data Management Coordinating Center Gateway
Database to dbGaP, the complete long read sequencing data is not immediately available.
Aggregate short read genome sequencing data on all UDN participants can be accessed via
dbGaP Study Accession: phs001232.v5.p2.

## DATA ACCESS

The data on HG002 Genome in a Bottle sample is available publicly and referenced in the Methods section. The Ilumina complete long read sequencing data will be available in dbGaP in accordance with Undiagnosed Diseases Network data sharing policies. As data is deposited from the Undiagnosed Disease Network Data Management Coordinating Center Gateway Database to dbGaP, the complete long read sequencing data is not immediately available. Aggregate short read genome sequencing data on all UDN participants can be accessed via dbGaP Study Accession: phs001232.v5.p2.

## ACKNOWLEDGEMENTS

<u>**Members of the Undiagnosed Diseases Network**</u>

Maria T. Acosta, David R. Adams, Ben Afzali, Ali Al-Beshri, Eric Allenspach, Aimee Allworth, Raquel L. Alvarez, Justin Alvey, Ashley Andrews, Euan A. Ashley, Carlos A. Bacino, Guney Bademci, Ashok Balasubramanyam, Dustin Baldridge, Erin Baldwin, Jim Bale, Michael Bamshad, Deborah Barbouth, Pinar Bayrak-Toydemir, Anita Beck, Alan H. Beggs, Edward Behrens, Gill Bejerano, Hugo J. Bellen, Jimmy Bennett, Jonathan A. Bernstein, Gerard T. Berry, Anna Bican, Stephanie Bivona, Elizabeth Blue, John Bohnsack, Devon Bonner, Nicholas Borja, Lorenzo Botto, Lauren C. Briere, Elizabeth A. Burke, Lindsay C. Burrage, Manish J. Butte, Peter Byers, William E. Byrd, Kaitlin Callaway, John Carey, George Carvalho, Thomas Cassini, Sirisak Chanprasert, Hsiao-Tuan Chao, Ivan Chinn, Gary D. Clark, Terra R. Coakley, Laurel A. Cobban, Joy D. Cogan, Matthew Coggins, F. Sessions Cole, Brian Corner, Rosario I. Corona, William J. Craigen, Andrew B. Crouse, Vishnu Cuddapah, Precilla D’Souza, Hongzheng Dai, Kahlen Darr, Surendra Dasari, Joie Davis, Margaret Delgado, Esteban C. Dell’Angelica, Katrina Dipple, Daniel Doherty, Naghmeh Dorrani, Jessica Douglas, Emilie D. Douine, Dawn Earl, Lisa T. Emrick, Christine M. Eng, Cecilia Esteves, Kimberly Ezell, Elizabeth L. Fieg, Paul G. Fisher, Brent L. Fogel, Jiayu Fu, William A. Gahl, Rebecca Ganetzky, Emily Glanton, Ian Glass, Page C. Goddard, Joanna M. Gonzalez, Andrea Gropman, Meghan C. Halley, Rizwan Hamid, Neal Hanchard, Kelly Hassey, Nichole Hayes, Frances High, Anne Hing, Fuki M. Hisama, Ingrid A. Holm, Jason Hom, Martha Horike-Pyne, Alden Huang, Yan Huang, Anna Hurst, Wendy Introne, Gail P. Jarvik, Suman Jayadev, Orpa Jean-Marie, Vaidehi Jobanputra, Oguz Kanca, Yigit Karasozen, Shamika Ketkar, Dana Kiley, Gonench Kilich, Eric Klee, Shilpa N. Kobren, Isaac S. Kohane, Jennefer N. Kohler, Bruce Korf, Susan Korrick, Deborah Krakow, Elijah Kravets, Seema R. Lalani, Christina Lam, Brendan C. Lanpher, Ian R. Lanza, Kumarie Latchman, Kimberly LeBlanc, Brendan H. Lee, Kathleen A. Leppig, Richard A. Lewis, Pengfei Liu, Nicola Longo, Joseph Loscalzo, Richard L. Maas, Ellen F. Macnamara, Calum A. MacRae, Valerie V. Maduro, Audrey Stephannie Maghiro, Rachel Mahoney, May Christine V. Malicdan, Rong Mao, Ronit Marom, Gabor Marth, Beth A. Martin, Martin G. Martin, Julian A. Martínez-Agosto, Shruti Marwaha, Allyn McConkie-Rosell, Ashley McMinn, Matthew Might, Mohamad Mikati, Danny Miller, Ghayda Mirzaa, Breanna Mitchell, Paolo Moretti, Marie Morimoto, John J. Mulvihill, Lindsay Mulvihill, Mariko Nakano-Okuno, Stanley F. Nelson, Serena Neumann, Dargie Nitsuh, Donna Novacic, Devin Oglesbee, James P. Orengo, Laura Pace, Stephen Pak, J. Carl Pallais, Neil H. Parker, LéShon Peart, Leoyklang Petcharet, John A. Phillips III, Filippo Pinto e Vairo, Jennifer E. Posey, Lorraine Potocki, Barbara N. Pusey Swerdzewski, Aaron Quinlan, Daniel J. Rader, Ramakrishnan Rajagopalan, Deepak A. Rao, Anna Raper, Wendy Raskind, Adriana Rebelo, Chloe M. Reuter, Lynette Rives, Lance H. Rodan, Martin Rodriguez, Jill A. Rosenfeld, Elizabeth Rosenthal, Francis Rossignol, Maura Ruzhnikov, Marla Sabaii, Jacinda B. Sampson, Timothy Schedl, Lisa Schimmenti, Kelly Schoch, Daryl A. Scott, Elaine Seto, Vandana Shashi, Emily Shelkowitz, Sam Sheppeard, Jimann Shin, Edwin K. Silverman, Giorgio Sirugo, Kathy Sisco, Tammi Skelton, Cara Skraban, Carson A. Smith, Kevin S. Smith, Lilianna Solnica-Krezel, Ben Solomon, Rebecca C. Spillmann, Andrew Stergachis, Joan M. Stoler, Kathleen Sullivan, Shamil R. Sunyaev, Shirley Sutton, David A. Sweetser, Virginia Sybert, Holly K. Tabor, Queenie Tan, Arjun Tarakad, Herman Taylor, Mustafa Tekin, Willa Thorson, Cynthia J. Tifft, Camilo Toro, Alyssa A. Tran, Rachel A. Ungar, Adeline Vanderver, Matt Velinder, Dave Viskochil, Tiphanie P. Vogel, Colleen E. Wahl, Melissa Walker, Nicole M. Walley, Jennifer Wambach, Michael F. Wangler, Patricia A. Ward, Daniel Wegner, Monika Weisz Hubshman, Mark Wener, Tara Wenger, Monte Westerfield, Matthew T. Wheeler, Jordan Whitlock, Lynne A. Wolfe, Heidi Wood, Kim Worley, Shinya Yamamoto, Zhe Zhang, Stephan Zuchner

