## Supplemental Information for "Clinical application of Complete Long Read genome sequencing identifies a 16kb intragenic duplication in EHMT1 in a patient with suspected Kleefstra syndrome"

Gorzynski et al.

**Supplemental Figure 1.** Phasing; Comparative analysis of ONT, PacBio, and ICLR technologies.

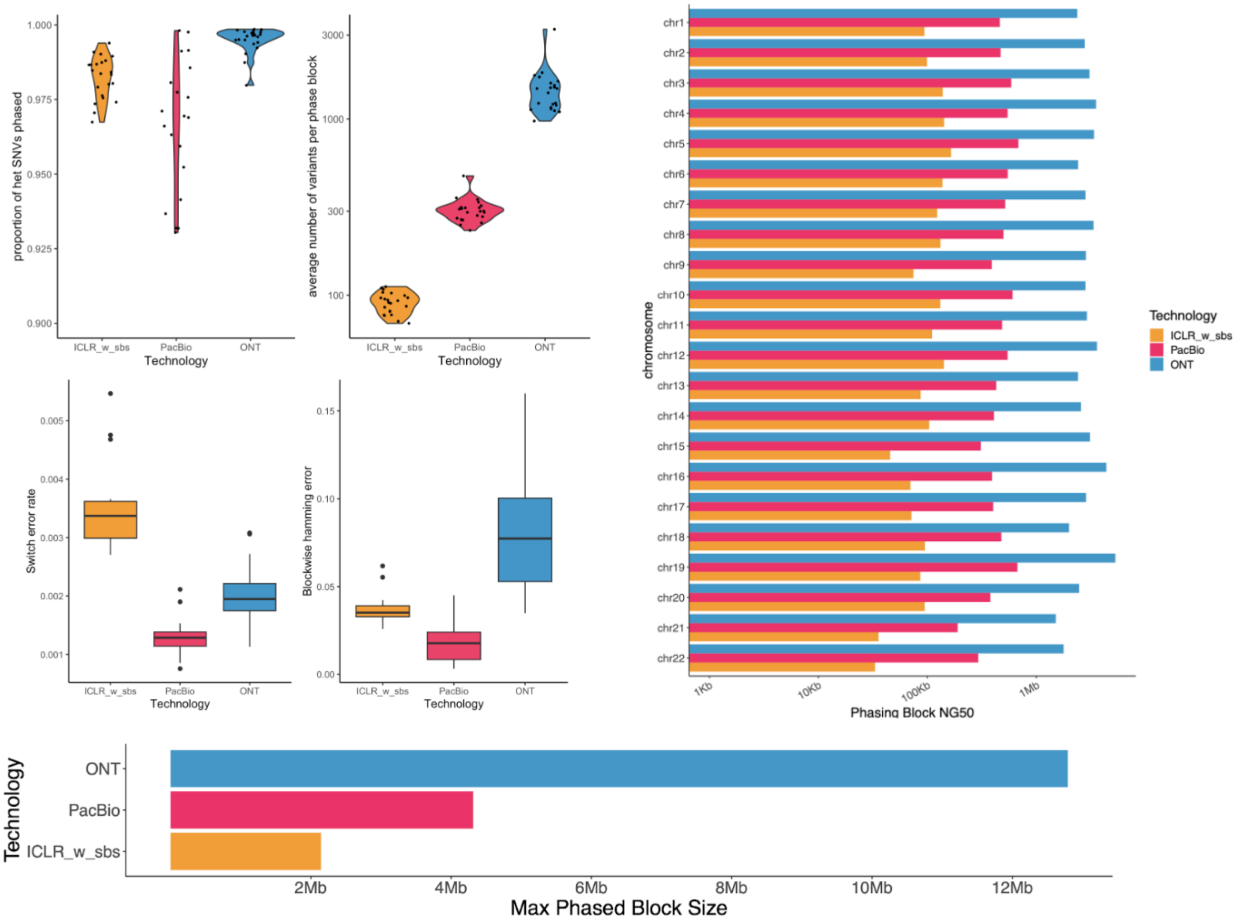

**Supplemental Figure 2.** Metrics including (A) mean coverage and (B) bases mapped (in Gigabases) are reported on the 8 ICLR bams. Bases mapped (cigar) are the number of mapped bases filtered by the CIGAR string corresponding to the read they belong to. Only alignment matches(M), inserts(I), sequence matches(=) and sequence mismatches(X) are counted. Total number of (C) SNVs and (D) SVs called are reported on the combined vcfs from ICLR and sbs (generated from short read sequencing). Each dot indicates an individual sample.

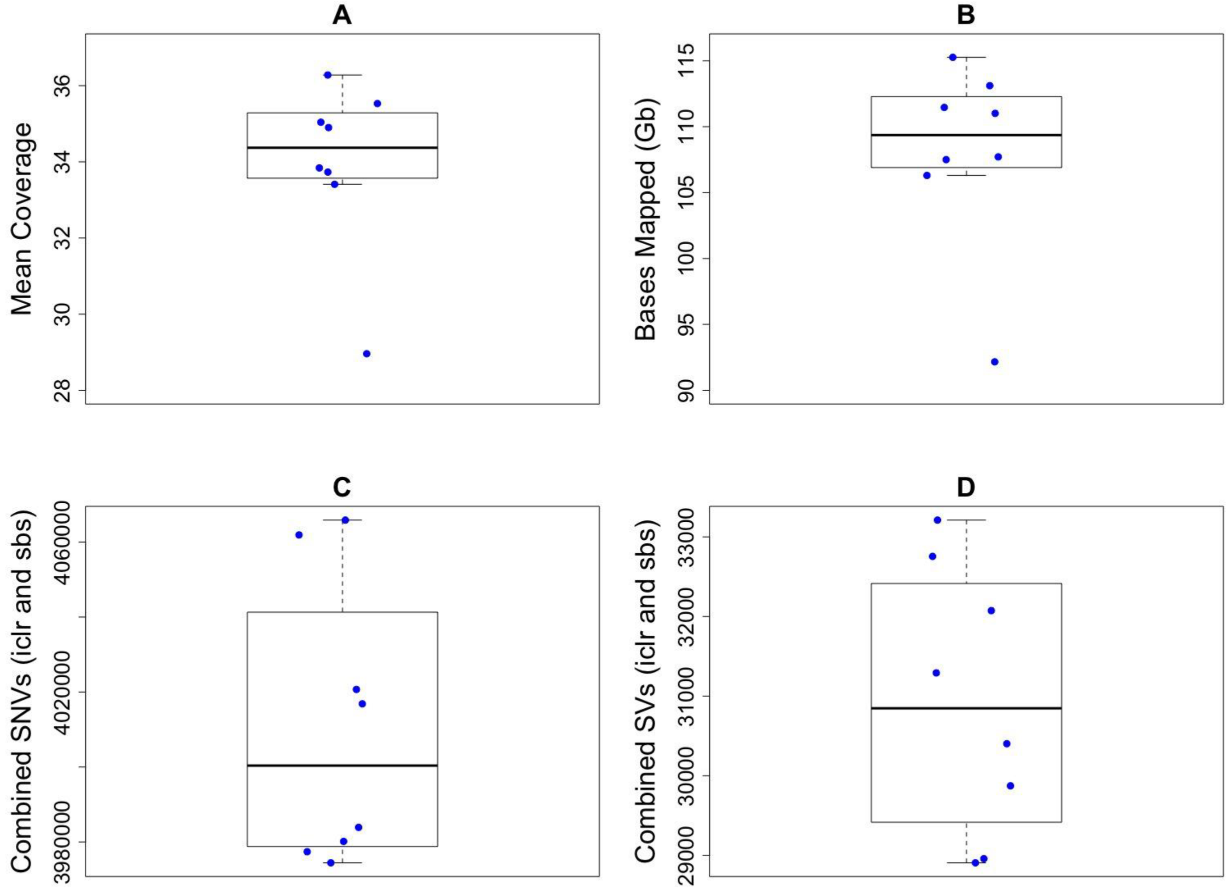

**Supplemental Table 1.** Metrics including N50, mean coverage and bases mapped (in Gigabases) are reported on the 8 ICLR bams. Bases mapped (cigar) are the number of mapped bases filtered by the CIGAR string corresponding to the read they belong to. Only alignment matches(M), inserts(I), sequence matches(=) and sequence mismatches(X) are counted. Total number of SNVs and SVs called are reported on the combined vcfs from ICLR and SBS (sequencing by synthesis, Illumina's short read sequencing).

| Sample | Type | N50 | Mean coverage :ICLR | Mean coverage: combined _iclr_sbs | Bases mapped (Gb) | # SNVs combined _iclr_sbs | # SVs combined _iclr_sbs |
|---|---|---|---|---|---|---|---|
| P1 | EDTA | 6351 | 34.90 | 72.07 | 111.01 | 3977406 | 29874 |
| P2 | EDTA | 6538 | 35.04 | 84.13 | 111.46 | 4061907 | 31292 |
| P3 | EDTA | 6626 | 36.28 | 76.01 | 115.26 | 3983893 | 32074 |
| P4 | PaxGene | 6394 | 33.84 | 86.30 | 107.71 | 3980156 | 32755 |
| P5 | PaxGene | 6663 | 33.41 | 87.60 | 106.30 | 4016854 | 30403 |
| P6 | Fibroblast | 6625 | 35.53 | 80.55 | 113.11 | 3974445 | 28959 |
| P7 | Fibroblast | 6733 | 28.96 | 78.74 | 92.16 | 4020695 | 33211 |
| HG002 | Fibroblast | 6588 | 33.73 | NA | 107.50 | 4065868 | 28907 |

**Supplemental Table 2**. Clinical genome sequencing details for ICLR samples.

| CLR_ID | SR-GS seq lab | Sequencing Platform |
|---|---|---|
| P1 | Baylor Genetics | Illumina HiSeq X |
| P2 | Baylor Genetics | Illumina HiSeq X |
| P3 | Baylor Genetics | Illumina HiSeq X |
| P4 | Baylor Genetics | Illumina HiSeq X |
| P5 | Baylor Genetics | Illumina HiSeq X |
| P6 | Hudson Alpha | Illumina NovaSeq |
| P7 | Hudson Alpha | Illumina NovaSeq |
| HG002 | n/a | n/a |

**Supplemental Table 3**. Clinical correlation of P7 with reported Kleefstra syndrome phenotype.

| **Characteristic** | **Typical KS phenotype**(Ciaccio et al. 2018) | **P7** |
|---|---|---|
| Microcephaly | 30-80% | + |
| Ear abnormalities | 45-80% | + overfolded helices |
| Dental anomalies | 10-15% | + delayed eruption of teeth |
| hypotonia | 60-80% | + in childhood |
| Psychomotor delay/intellectual disability | 100% | + |
| Autism | 30-75% | |
| Behavioral problems | 65-70% | |
| Sleep disorder | 20-50% | |
| epilepsy | 20-50% | |
| Brain imaging anomalies | 50-60% | |
| Cardiovascular anomalies | 40-45% | + atrial fibrillation |
| Renal issues | 15-30% | |
| Genital anomalies | 45-50% | |
| hernia | 15-20% | + inguinal hernia |
| Skeletal anomalies | 30-50% | + thoracic kyphosis, syndactyly |
| Hearing impairment | 20-30% | + mild bilateral hearing loss |
| Ocular anomalies | 45% | |
| obesity | 30-40% | + truncal obesity |
| Dysmorphic features | Most | + |
